# Transcriptomic clustering of critically ill COVID-19 patients

**DOI:** 10.1101/2022.03.01.22271576

**Authors:** Cecilia López-Martínez, Paula Martín-Vicente, Juan Gómez de Oña, Inés López-Alonso, Helena Gil-Peña, Elías Cuesta-Llavona, Margarita Fernández-Rodríguez, Irene Crespo, Estefanía Salgado del Riego, Raquel Rodríguez-García, Diego Parra, Javier Fernández, Javier Rodríguez-Carrio, Alberto Dávalos, Luis A Chapado, Eliecer Coto, Guillermo M Albaiceta, Laura Amado-Rodríguez

**Author notes:** GMA and LAR share last authorship. Authors for correspondence: Guillermo M Albaiceta, Unidad de Cuidados Intensivos Cardiológicos, Hospital Universitario Central de Asturias, Avenida del Hospital Universitario s/n, 33011 Oviedo. Spain., Laura Amado-Rodríguez, Unidad de Cuidados Intensivos Cardiológicos, Hospital Universitario Central de Asturias, Avenida del Hospital Universitario s/n, 33011 Oviedo. Spain.

## Abstract

Infections caused by SARS-CoV-2 may cause a severe disease, termed COVID-19, with significant mortality. Host responses to this infection, mainly in terms of systemic inflammation, have emerged as key pathogenetic mechanisms, and their modulation is the only therapeutic strategy that has shown a mortality benefit. Herein, we used peripheral blood transcriptomes of critically-ill COVID-19 patients obtained at admission in an Intensive Care Unit (ICU), to identify two transcriptomic clusters characterized by expression of either interferon-related or immune checkpoint genes, respectively. These profiles have different ICU outcome, in spite of no major clinical differences at ICU admission. A transcriptomic signature was used to identify these clusters in an external validation cohort, yielding similar results. These findings reveal different underlying pathogenetic mechanisms and illustrate the potential of transcriptomics to identify patient endotypes in severe COVID-19, aimed to ultimately personalize their therapies.

---

Infections caused by SARS-CoV-2 have a wide range of severity, from asymptomatic to life-threatening cases. The most severe forms of Coronavirus-induced disease (termed COVID-19) ^1^ lead to respiratory failure fulfilling the acute respiratory distress syndrome (ARDS) criteria ^2^. These critically ill patients often require mechanical ventilation and supportive therapy in an intensive care unit (ICU) and show mortality rates that range from 12 to 91% depending on patient and hospital factors ^3^.

Local and systemic inflammation are key pathogenetic mechanisms in severe COVID-19 ^4^. Viral infection triggers a host response that involves not only anti-viral mechanisms, such as release of interferons, but may also activate a systemic, non-specific inflammatory response that has been related to multiple organ failure and death ^5^. The only treatments that have shown a survival benefit in critically-ill COVID-19 patients aim to modulate this inflammatory response ^6^. However, it has been suggested that these treatments do not benefit patients with less severe forms of the disease or with only a mild activation of inflammation ^7,8^.

There is increasing evidence that ARDS patients show different clinical features or systemic responses to severe diseases (phenotypes and endotypes respectively) ^9^. Although the underlying causes responsible for this heterogeneity are not fully understood, clinical data showing different outcomes in response to a given treatment suggest that pathogenetic mechanisms may be different ^10^. Therefore, identification of patient pheno/endotypes may be relevant not only for risk stratification, but also to design specific, personalized therapies in the ICU.

Several studies have tried to identify severe COVID-19 phenotypes using clinical data, yielding sometimes conflicting results. Although translation of the previously identified ARDS phenotypes to COVID-19 showed two groups of patients with different responses to steroid therapy ^11^, other studies failed to identify clear groups of patients using clinical data at admission ^12^.

Transcriptomic profiling after sequencing of whole blood RNA may be useful to identify groups of critically-ill patients with different underlying pathogenetic mechanisms ^13–15^. In addition, preliminary results suggest that micro-RNA (miRNA) expression could also play a role in this setting ^16^. We hypothesized that clustering of COVID-19 patients using transcriptomics at ICU admission could help to identify subgroups with different pathogenesis. To test this hypothesis, we prospectively sequenced peripheral blood RNA and serum miRNA at ICU admission in a cohort of COVID-19 patients, applied an unbiased clustering algorithm and compared gene expression clinical data and outcomes in the identified subgroups. Finally, we validated our findings in an external cohort.

## Methods

### Study design

This prospective observational study was reviewed and approved by the regional ethics committee (Comité de Ética de la Investigación Clínica del Principado de Asturias, ref 2020.188). Informed consent was obtained from each patient’s next of kin. Fifty-six consecutive patients admitted to one of the participant ICUs at Hospital Universitario Central de Asturias (Oviedo, Spain) from April to December 2020 were included in the study. Inclusion criteria were ICU admission and PCR-confirmed COVID-19. Exclusion criteria were age<18, any condition that could explain the respiratory failure other than COVID-19, do-not-resuscitate orders or terminal status, refusal to participate or severe comorbidities that may alter the systemic response (immunosuppression, history of organ transplantation, disseminated neoplasms). All patients were managed following a standardized written clinical protocol.

### Sample acquisition and processing

After inclusion, two samples of peripheral blood were drawn in the first 72 hours after ICU admission. One sample was collected in Tempus Blood RNA tubes (Thermo Fisher) to facilitate cell lysis, precipitate RNA and prevent its degradation. The other sample was immediately centrifuged to obtain serum and mixed with TRI reagent for serum RNA precipitation. These tubes were stored at -80°C until processing. Whole blood RNA was extracted by isopropanol precipitation and sequenced in an Ion S5 GeneStudio sequencer using AmpliSeq Transcriptome Human Gene Expression kits that amplify all the canonical human transcripts. Details on RNA extraction and sequencing have been provided elsewhere ^8^. FASTQ files containing RNA sequences were pseudoaligned using a reference transcriptome (http://refgenomes.databio.org) and *salmon* software ^17^ to obtain transcript counts.

Total serum RNA was extracted using miRNEasy kit (Qiagen), following manufacturer’s instructions, and miRNAs isolated and sequenced at BGI Genomics (Wuhan, China). miRNA readouts were mapped using *bowtie2* ^18^, with an index built using the hg38 human reference genome. Quantification of sequenced miRNAs was performed using *miRDeep2* ^19^ with reference human mature and hairpin miRNA sequences downloaded from miRBase (release 22, https://www.mirbase.org).

### Clustering

Clustering of RNA samples was performed following a previously described protocol ^20^. Briefly, log_2_-transformed gene expression data (expressed as transcripts per million reads) were filtered to keep the 5% of features with the largest variance. Clusters were built based on Euclidean distances following the Ward clustering algorithm. Cluster p-values, indicating how strong the cluster is supported by the data, were calculated by multiscale bootstrap resampling using the *pvclust* package ^21^ for R.

### Analysis of differentially expressed genes

Gene raw counts obtained after pseudoalignment were compared between clusters using *DESeq2* ^22^. Log_2_ fold change for each gene between variants and the corresponding adjusted p-value (corrected using a false discovery rate of 0.05) were calculated. Genes with an absolute log_2_ fold change above 2 and an adjusted p-value lower than 0.01 were used for Gene Set Enrichment Analysis (GSEA) using the *clusterProfiler* R package ^23^.

A correlation analysis was performed in genes annotated to a Gene Ontology category involved in interferon pathway. Correlation coefficients between each gene pair were transformed to z-scores and the p-values for each comparison calculated using the *DGCA* package for R ^24^. Genes with opposite correlations in each cluster were selected and the networks defined by their significant correlations traced.

Differentially expressed genes between clusters were also matched with the c-miRNAs expressed for each group using the *MicroRNA Target Filter* tool from *Ingenuity Pathway Analysis* (Qiagen Digital Insights), to identify predicted interactions. Intersected mRNA and miRNA datasets were filtered to explicitly pair opposed and reciprocal expression changes. Only experimentally observed predictions were considered. Key mRNA-miRNA relationships identified were overlayed onto the networks of interest to explore the predicted functionality in our datasets. Pathways related to humoral, and T and B cellular immune responses were selected as relevant. miRNAs with <3 targeted mRNAs were filtered out from the network.

### Clinical data

Demographics and comorbidities were collected at ICU admission (day 1). Data on gas exchange, respiratory support, hemodynamics, received treatments and results from routine laboratory analyses were prospectively collected at days 1 and 7 after ICU admission. Patients were followed up to ICU discharge. During this period, duration of ventilatory support and vital status were collected for outcome analysis.

### Circulating cell populations

Proportions of transcriptionally active circulating cells in each sample were estimated using *Immunostates* ^25^, a previously published deconvolution algorithm. From the original reference matrix, cell populations not commonly identified in peripheral blood (Mast cells and macrophages) were removed. Using this modified reference matrix containing expression of 318 genes for 16 different blood cell types, the percentage of each one of these types was estimated from the bulk RNAseq.

### Validation

To validate our results in an external cohort, we used a publicly available dataset of 50 transcriptomes from critically-ill COVID-19 patients ^26^. Sample acquisition was performed at enrolment. Clinical data and gene counts were downloaded from Gene Expression Omnibus (https://www.ncbi.nlm.nih.gov/geo/, accession number GSE157103). First, we identified differentially expressed genes that best discriminate between clusters in our data, as those with an Area Under the Receiver Operating Characteristic curve (AUROC) above 0.95. A transcriptomic score was calculated as the geometric mean of these genes, and the AUROC for this score determined and a threshold between clusters was defined. Finally, the same transcriptomic score was calculated in the validation cohort, and each sample assigned to one cluster using the previously established threshold. Clinical data (age, sex, APACHE-II score) and outcomes (ventilator-free days -VFDs-at ICU Day 28) were compared between clusters.

### Statistical analysis

Data are expressed as median and interquartile range. Missing data were not imputed. Differences between clusters were assessed using two-tailed Wilcoxon or chi-square tests (for quantitative and qualitative data respectively). For survival analysis, patients were followed up to ICU discharge, with ICU discharge alive and spontaneously breathing being the main outcome measurement. Differences in this outcome between clusters were assessed using a competing risk model as previously described ^8^, and hazard ratio for the main outcome, with the corresponding 95% confidence interval, was calculated. All the analyses were performed using R v4.1.1 ^27^ and packages *ggplot2* ^28^, pROC ^29^ and *survival* ^30^, in addition to those previously cited. All the code and raw data can be found at https://github.com/Crit-Lab/COVID_clustering.

## Results

### Patient clustering

Peripheral gene expression was sequenced in 56 consecutive critically-ill patients (20% female, age 68 [61 - 75] year) admitted to one of the participant ICUs. Amongst 16903 genes counted, 1727 were used for hierarchical clustering (Figure 1A). The two main branches of the obtained clustering tree showed the highest p values (Figure 1B and supplementary Figure 1). Therefore, the sample was divided in two mutually exclusive groups, termed COVID-19 transcriptomic profiles (CTP) 1 and 2. Bidimensional representation of the study population using a UMAP algorithm confirmed the separation of the two clusters (Figure 1C). Supplementary figure 2 shows a heatmap with the expression of the genes used for clustering.

**Figure 1.**
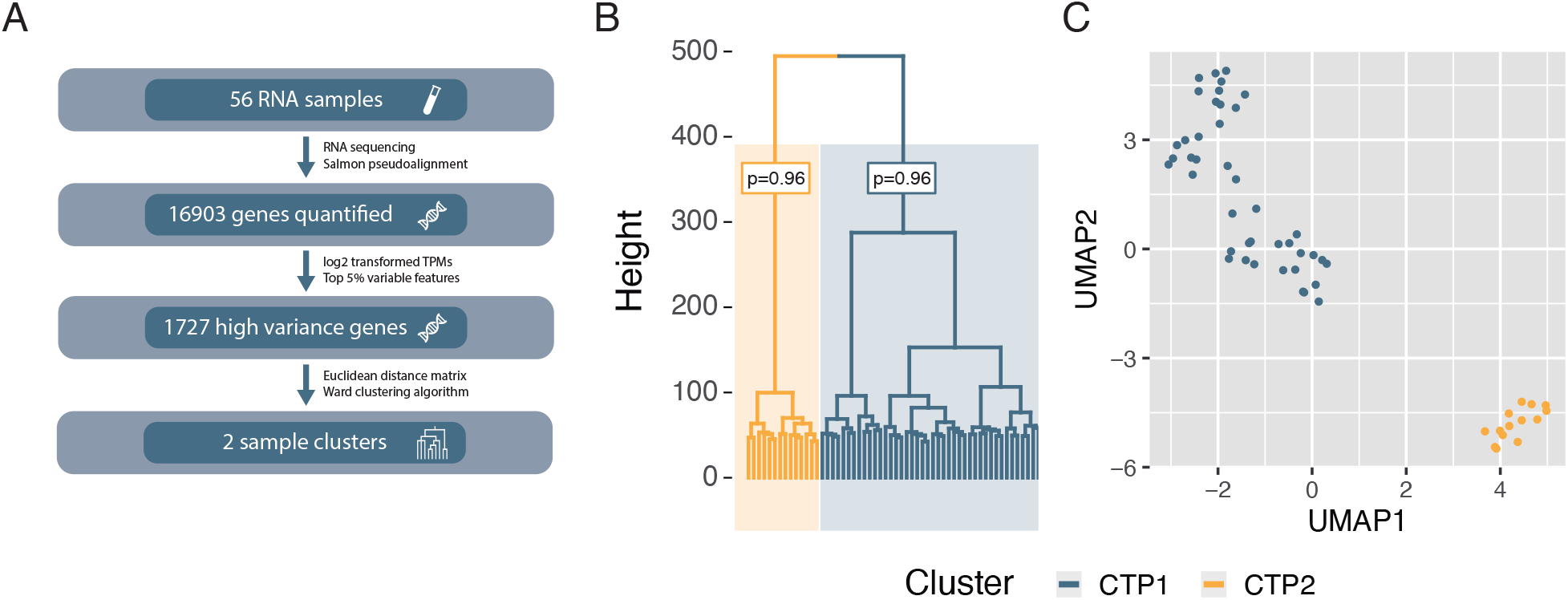
Patient clustering. A: Clustering strategy based on peripheral blood RNAseq, using the 5% genes with the highest variance among samples. B: Hierarchical clustering tree, showing the p-values (corresponding to the alternative hypothesis that the cluster does not exist) of the two main clusters. C: Uniform manifold approximation and projection (UMAP) showing a bidimensional representation of all the samples and clusters. TPM: Transcripts per million reads.

### Differences between transcriptomic profiles

Then we assessed the overall differences in gene expression. Using an adjusted p-value cut-off point of 0.01, there were 9700 differentially expressed genes (Supplementary file 1), with 3640 having an absolute log_2_ fold change above 2 (Figure 2A). Interestingly, most of these genes were downregulated in CTP2. Then, GSEA was used to identify the molecular pathways involving these differentially expressed genes. One hundred and ten biological processes with significant differences between clusters were identified (Supplementary Figure 3). Among these, several categories related to the interferon-mediated response and lymphocyte activation were identified (Figure 2B), and participating genes were plotted (Figure 2C-E). Patients included in CTP1 showed an enrichment of several interferon genes, linked to the activation of a number of immune populations related to innate and adaptative responses (Figure 2C), whereas CTP2 was enriched in genes involved in B-cell receptor signaling (Figure 2D) and regulatory T-cell differentiation (Figure 2E).

**Figure 2.**
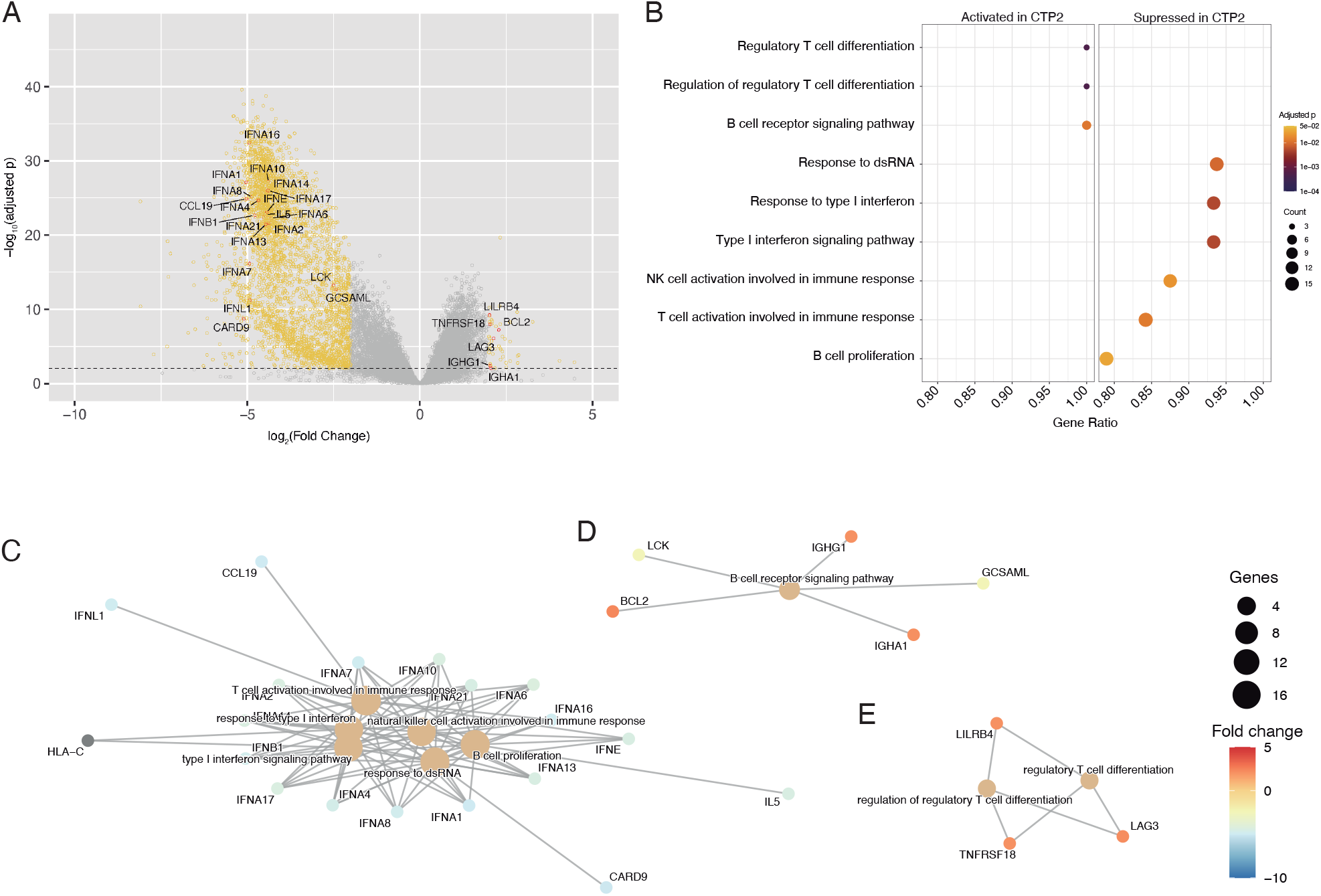
Differentially expressed genes between COVID transcriptomic clusters (CTP). A: Volcano plot showing fold-change for each gene and their significance level. Genes with an adjusted p-value lower than 0.01 and an absolute log_2_ fold change above 2 are colored in orange. Differentially expressed genes included in interferon-dependent pathway are labelled. B: Enrichment of Gene Ontology categories related to Interferon signaling in COVID Transcriptomic Profile 2 (CTP2, n=14 compared to CTP1, n=42). C-E: Gene functional networks with differential expression between clusters, involving Interferon-dependent lymphoid activation (C), B-cell receptor signaling (D) and regulatory T-cell differentiation (E).

In addition to these quantitative changes in expression of interferon-related genes, we explored the existence of qualitative differences between clusters. We calculated the linear correlation coefficients among the 145 genes included in the Gene ontology categories involving interferon signaling in each cluster. There was a significant difference between the two correlation matrices (Figure 3, p<0.001 calculated using a Chi-square test), thus demonstrating differences in the orchestration/structure of IFN responses between groups. In addition, pairwise differences in correlation coefficients for each gene pair were assessed. Gene pairs with correlation coefficients with an adjusted p-value for their difference below 0.05 and opposite signs in each cluster were selected, and networks including these genes traced (Figure 3 and Supplementary Figure 4). These results suggest that both clusters have a qualitatively different activation of the interferon pathway, with some genes such as HSP90AB1 and JAK1 acting as hubs with opposite correlations. Of note, CTP1 was hallmarked by strong, positive correlations among effector IFN proteins, whereas this was not the case for CTP2.

**Figure 3.**
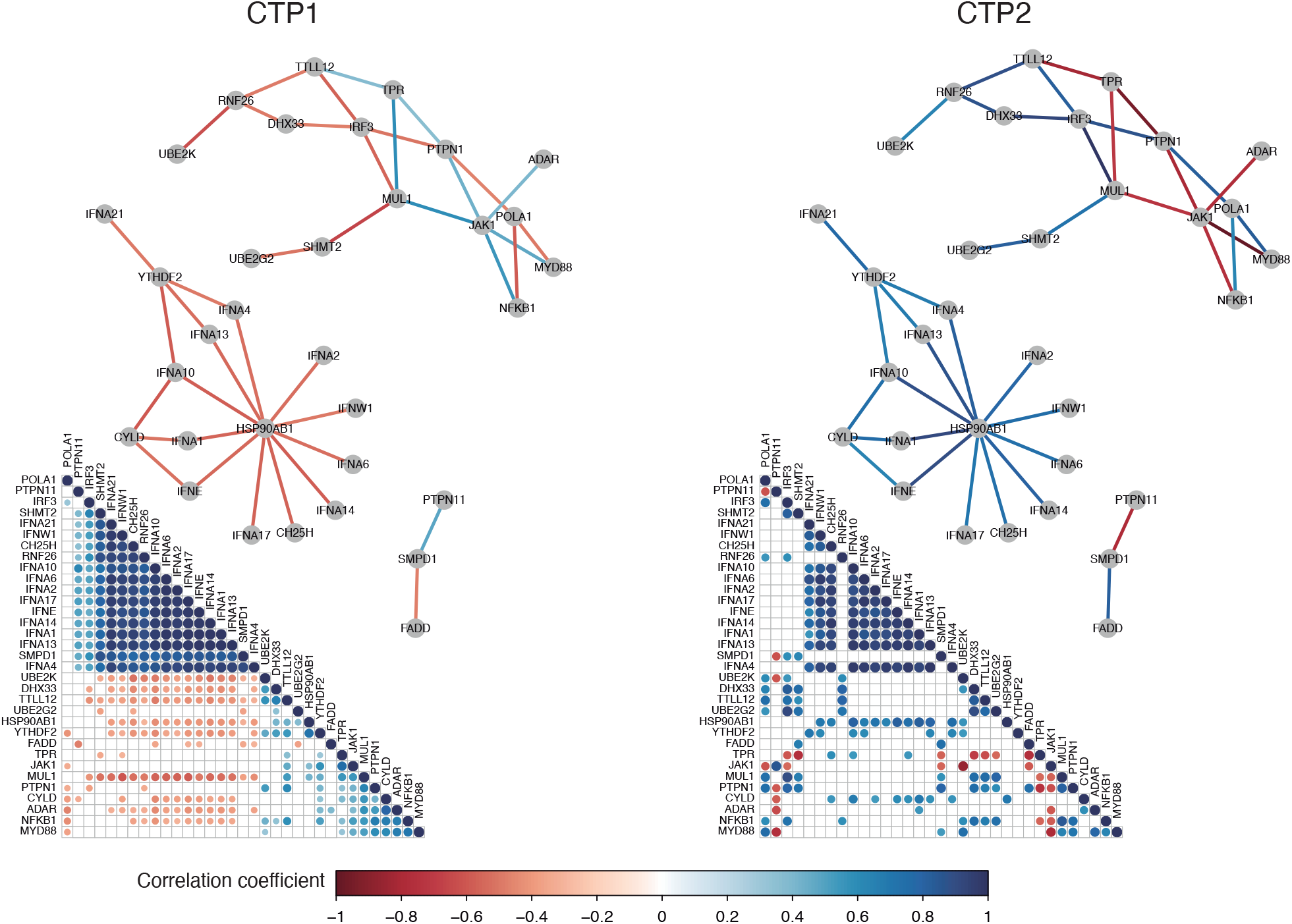
Correlation between genes included in Interferon-dependent pathways. Correlograms (bottom) and gene networks (top) showing correlations with opposite sign between genes in each COVID transcriptomic cluster (CTP, n=42 and 14 for CTP1 and 2 respectively). Only Pearson correlation coefficients with a P-value lower than 0.05 are shown.

### Differences in circulating cell populations

The previous results suggest that the identified clusters may have a different circulating lymphocyte profile. To further explore this finding, cell populations were estimated by deconvolution of RNAseq data. This analysis revealed a higher granulocyte proportion in patients assigned to CTP1, a lower proportion of lymphocytes and no differences in monocytes or NK cells (Figure 4A-D). Although no differences in absolute lymphocyte counts were found (645 [483 — 948] vs 730 [580 – 908] /mm3, p=0.71, Table 1), deconvolution and adjustment by total lymphocyte fraction revealed a higher proportion of CD4+ T cells (Figure 4E) and a lower proportion of CD8+ T cells (Figure 4F) and naïve B-cells (Figure 4G), with no differences in memory B-cells (Figure 4H) in this group, in line with the GSEA results. Detailed data on other cell populations can be found in the online supplement (supplementary figure 5).

**Figure 4.**
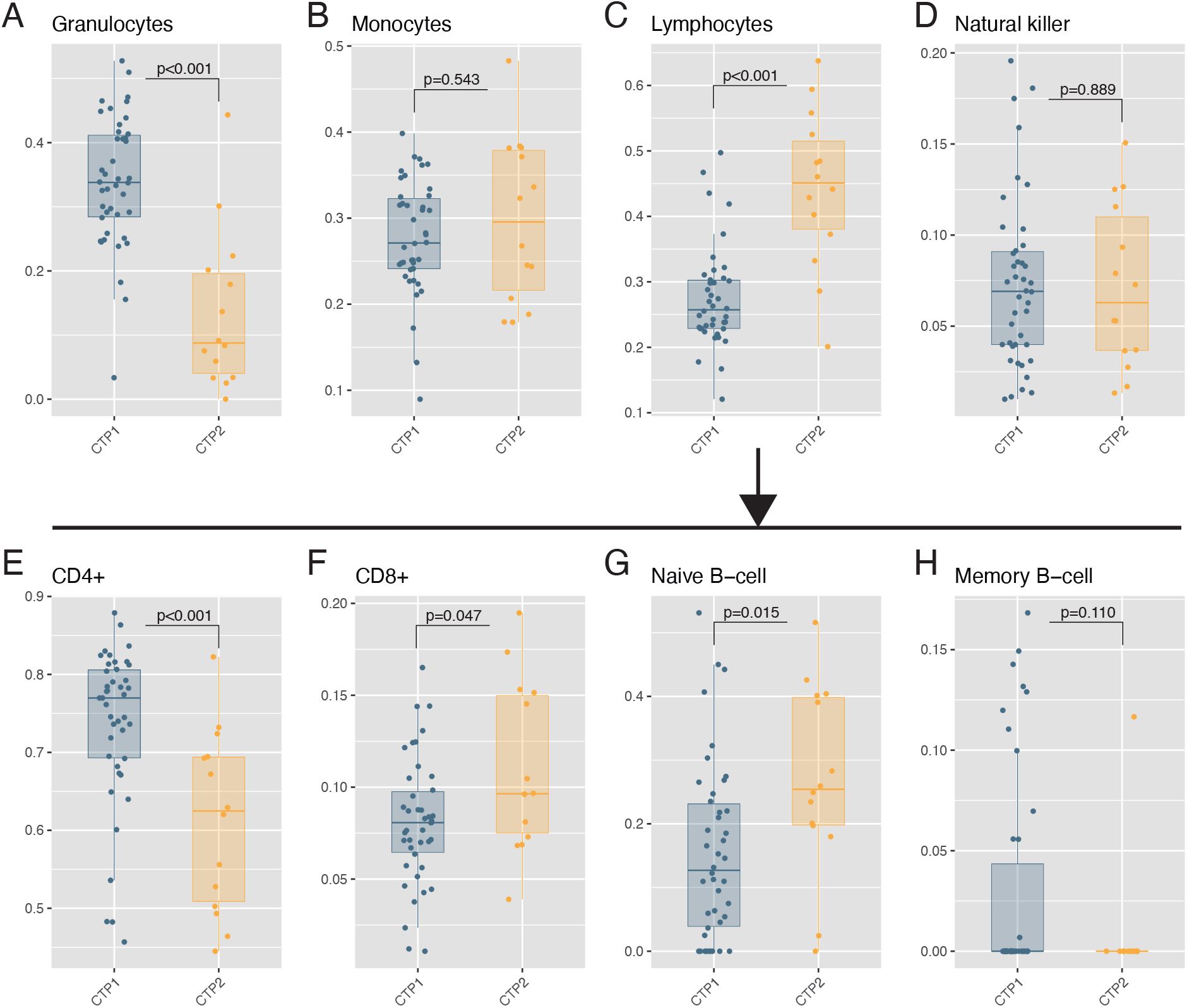
Estimated circulating cell populations. A-D: Proportions of blood cells were estimated from RNA-seq using a deconvolution algorithm. E-H: Lymphocyte subpopulations expressed as percentage of the absolute number of lymphocytes. Points represent individual patient data. In boxplots, bold line represents the median, lower and upper hinges correspond to the first and third quartiles (the 25th and 75th percentiles) and upper and lower whiskers extend from the hinge to the largest or smallest value no further than 1.5 times the interquartile range. P-values were calculated using a two-tailed Wilcoxon test.

**Table 1.** Clinical differences between COVID transcriptomic profiles (CTP). BMI: Body mass index. COPD: Chronic Obstructive Pulmonary Disease. APACHE-II: Acute Physiology and Chronic Health disease Classification System II. PBW: Predicted body weight (according to height). PEEP: Positive end-expiratory pressure. IL-6: Interleukin-6. Data are expressed as median (interquartile range) or count (percentage). P-values were calculated using a Wilcoxon test (quantitative data) or Chi-square test (proportions).

|  | CTP1 (n=42) | CTP2 (n=14) | p_value |
| --- | --- | --- | --- |
| Sex |  |  | 0.174 |
| Male | 36 (86%) | 9 (64%) |  |
| Female | 6 (14%) | 5 (36%) |  |
| Age (years) | 69 (63 - 75) | 63.5 (59 - 69) | 0.147 |
| BMI (Kg/m<sup>2</sup>) | 29 (25 - 33) | 29 (27 - 31) | 0.781 |
| Race |  |  | 0.582 |
| Caucasian | 38 (90%) | 14 (100%) |  |
| Black | 2 (5%) | 0 |  |
| Latino | 2 (5%) | 0 |  |
| Chronic kidney disease | 4 (10%) | 0 | 0.549 |
| COPD | 5 (12%) | 1 (7%) | 1 |
| Liver cirrhosis | 1 (2%) | 0 | 1 |
| Arterial hypertension | 26 (62%) | 6 (43%) | 0.35 |
| Diabetes | 9 (21%) | 3 (21%) | 1 |
| Dyslipemia | 18 (43%) | 6 (43%) | 1 |
| Day 1 |  |  |  |
| APACHE-II score | 18 (14 - 21) | 16 (13 - 17) | 0.120 |
| FiO<sub>2</sub> | 0.5 (0.4 - 0.6) | 0.45 (0.3 - 0.5) | 0.438 |
| PaO<sub>2</sub>/FiO<sub>2</sub> | 197 (157 - 245) | 188 (151 - 278) | 0.863 |
| PaCO<sub>2</sub> (mmHg) | 43 (39 - 47) | 41 (39 - 42) | 0.091 |
| Respiratory rate (/min) | 18 (16 - 21) | 18 (17 - 22) | 0.859 |
| pH | 7.37 (7.32 - 7.41) | 7.42 (7.36 - 7.43) | 0.099 |
| Lactate (mEq/L) | 1.3 (1.08 - 1.8) | 1.1 (0.9 - 1.2) | 0.040 |
| Tidal volume (ml) | 479 (455 - 504) | 500 (475 - 514) | 0.499 |
| Tidal volume / PBW (ml/Kg) | 7.5 (6.9 - 8.3) | 8 (7.5 - 8.7) | 0.239 |
| Plateau pressure (cmH<sub>2</sub>O) | 27 (24 - 29.75) | 25 (22 - 29) | 0.776 |
| PEEP (cmH<sub>2</sub>O) | 14 (12 - 15) | 12 (10 - 12) | 0.088 |
| Driving pressure (cmH<sub>2</sub>O) | 14 (11 - 15) | 15 (12 - 15) | 0.568 |
| Compliance (ml/cmH<sub>2</sub>O) | 36 (31 - 43) | 31 (29 - 44) | 0.697 |
| Creatinin (mg/dl) | 0.92 (0.68 - 1.23) | 0.71 (0.59 - 0.97) | 0.130 |
| Procalcitonin (ng/ml) | 0.23 (0.14 - 0.6) | 0.14 (0.13 - 0.27) | 0.250 |
| IL-6 (pg/ml) | 113 (54 - 276) | 164 (36 - 250) | 0.784 |
| Ferritin (ng/ml) | 1329 (968 - 1606) | 1673 (856 - 2182) | 0.576 |
| Leukocytes (/μl) | 9010 (6750 - 11825) | 5440 (4418 - 6453) | 0.002 |
| Lymphocytes (/μl) | 645 (482.5 - 948) | 730 (580 - 908) | 0.705 |
| D-dimer (ng/ml) | 1495 (842 - 3304) | 1084 (750 - 2126) | 0.501 |
| Days from hospital to ICU admission | 2 (0 - 3) | 2 (1 - 4) | 0.5 |
| Treatments during ICU stay |  |  |  |
| Mechanical ventilation | 38 (90%) | 11 (79%) | 0.484 |
| Prone ventilation | 23 (61%) | 8 (73%) | 0.981 |
| Neuromuscular blockade | 23 (61%) | 6 (55%) | 0.643 |
| Extracorporeal membrane oxygenation | 1 (3%) | 0 | 1 |
| Vasoactive drugs |  |  | 0.204 |
| None | 17 (40%) | 6 (43%) |  |
| One | 25 (60%) | 7 (50%) |  |
| Two or more | 0 | 1 (7%) |  |
| Steroid therapy | 19 (45%) | 5 (36%) | 0.755 |
| ICU evolution |  |  |  |
| IL-6 at day 7 | 54 (11 - 171) | 42 (16 - 130) | 0.713 |
| Ferritin at day 7 | 1100 (698 - 1504) | 1544 (805 - 1908) | 0.745 |
| D-dimer at day 7 | 2068 (1249 - 4586) | 1541 (988 - 3370) | 0.422 |
| Ventilator-free days at day 28 | 12 (0 - 19) | 19 (9 - 23) | 0.050 |

### Potential regulatory miRNAs

To identify miRNAs potentially related to the observed changes in RNA expression, we analyzed miRNA content using the *Mirna Target Filter* included in *Ingenuity Pathway Analysis*. After filtering by experimentally confirmed miRNA-gene relationships, and only opposed changes in miRNA/gene expression levels, 83 miRNAs targeting 608 genes were identified in our dataset. Given the observed differences in lymphocyte populations, we focused on miRNAs involved in humoral and cellular immune regulation (29 miRNAs and 151 genes). Paired miRNA-gene networks are depicted in Supplementary Figure 6 (104 downregulated genes/18 predicted upregulated miRNAs) and Figure 5 (47 upregulated genes/11 predicted downregulated miRNAs), with an overlay including differentially expressed genes between CTP1 and CTP2. miRNAs predicted to regulate expression of these genes were identified and compared (Figure 5B-H). Among these, only counts of miR-145a-5p and miR-181-5p were significatively lower in CTP2 (Figure 5C and 5D respectively).

**Figure 5.**
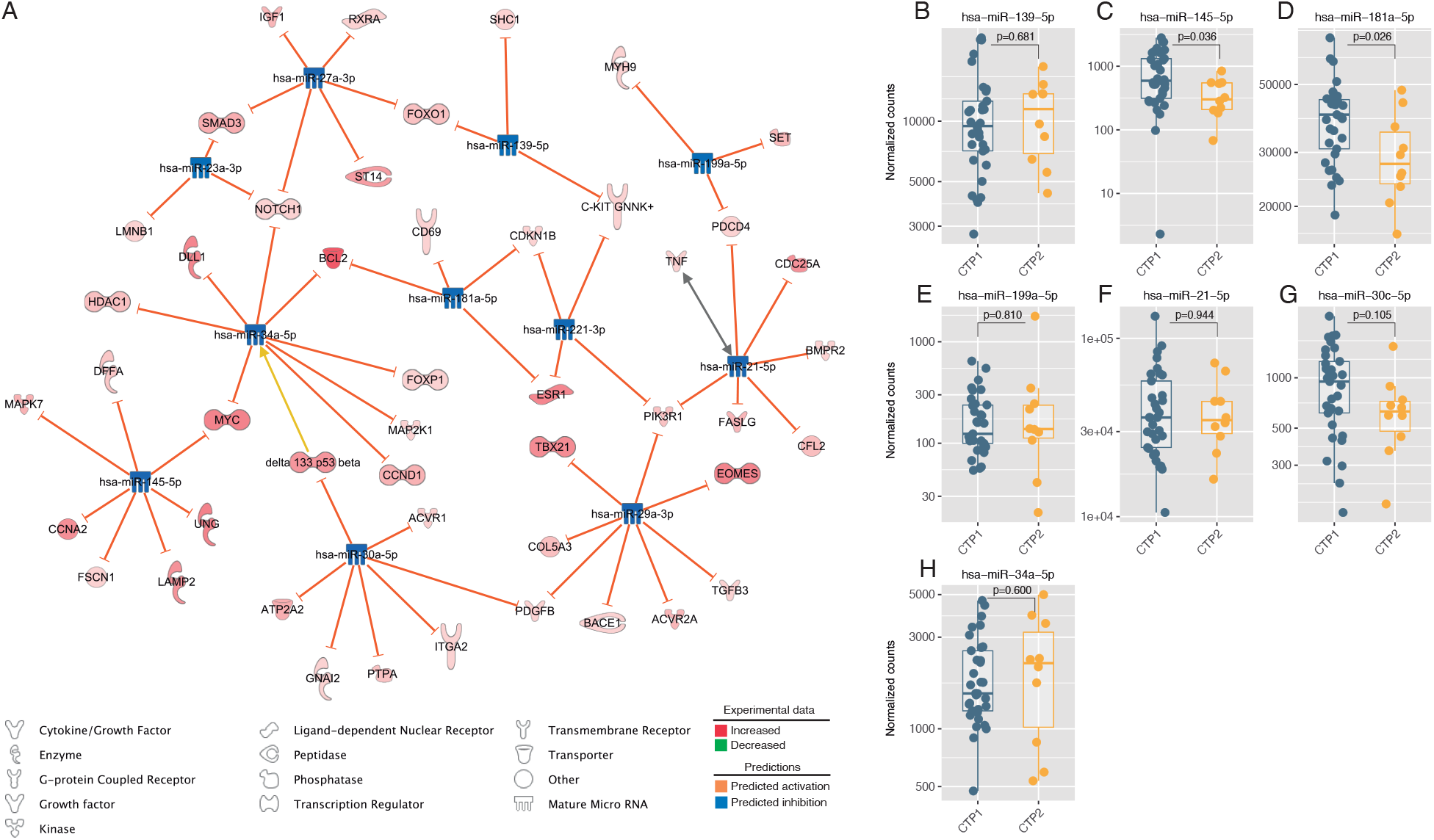
Regulation of gene expression by micro-RNAs. A: Micro-RNAs potentially regulating genes with increased differential expression were identified and a network built. B-H: Counts of hub micro-RNAs (defined as those regulating 3 or more differentially expressed genes) in serum. Points represent individual patient data. In boxplots, bold line represents the median, lower and upper hinges correspond to the first and third quartiles (the 25th and 75th percentiles) and upper and lower whiskers extend from the hinge to the largest or smallest value no further than 1.5 times the interquartile range. P-values were calculated using a two-tailed Wilcoxon test.

### Clinical differences and outcome

Clinical differences between clusters at ICU admission were studied (Table 1). There were no significant differences in demographic and clinical variables other than a higher leukocyte count in cluster CTP1, with no differences in lymphocyte counts. Patients assigned to CTP2 cluster showed more ventilator-free days during the first 28 days in ICU (Table 1). In the survival analysis, after adjusting for age, sex, and need for intubation during the ICU stay, assignation to CTP2 increased the probability of ICU discharge alive and spontaneously breathing (HR 2.00 [1.08 – 3.70], p=0.028, Figure 6).

**Figure 6.**
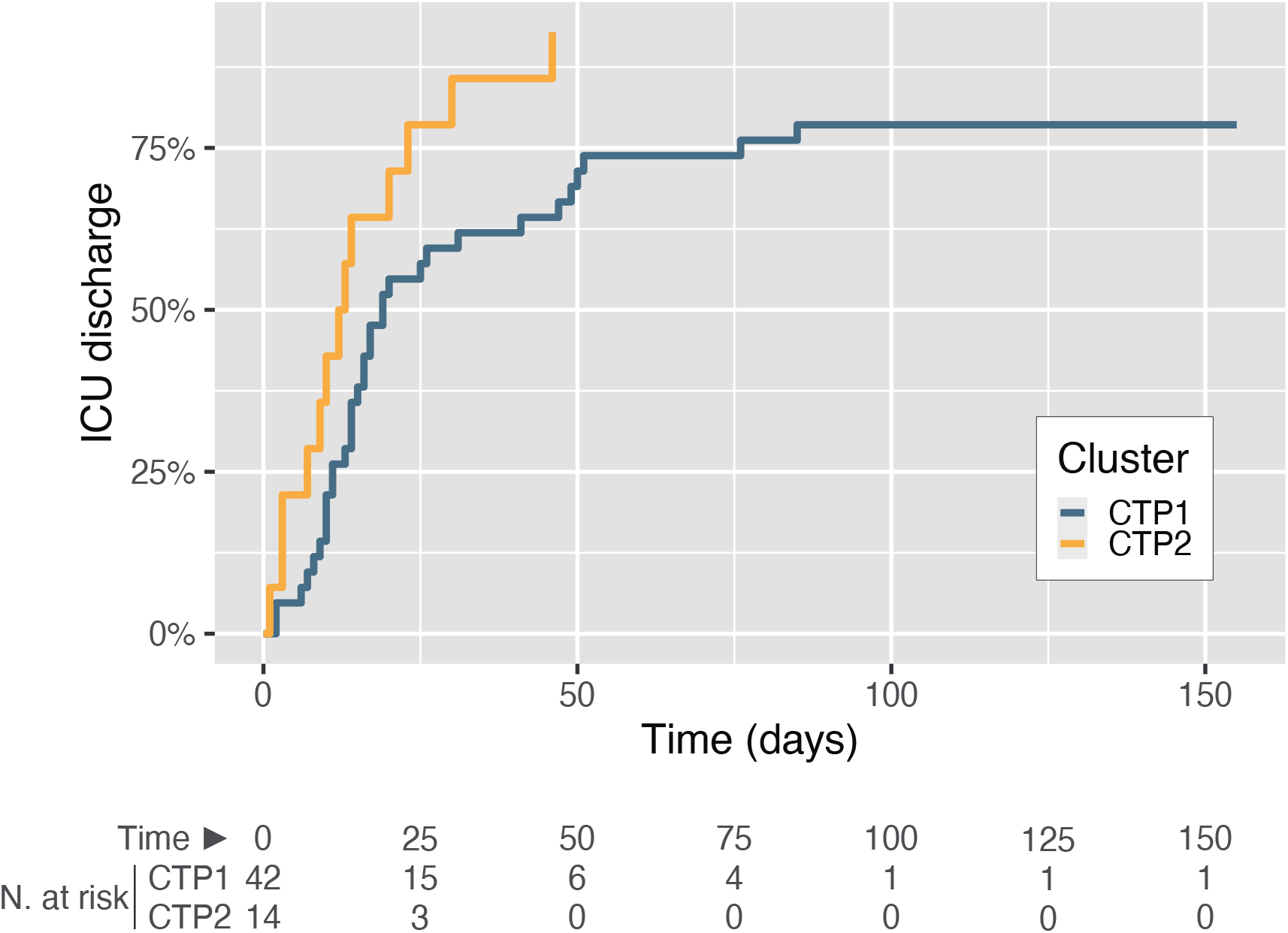
Intensive Care Unit (ICU) stay. Cumulative incidence of the main outcome (ICU discharge alive and spontaneously breathing), modelled using a competing risk model (with death as a competitive risk) and adjusted by age, sex and need for mechanical ventilation during the ICU stay.

### External validation

To apply our findings to an external cohort, we first developed a characteristic gene signature that allows assignation to one cluster using gene expression data. We focused on genes upregulated in CTP2, as they constitute a relatively small group, given the massive gene downregulation in this group. Among these 117 upregulated genes, 15 (BCL2, CARD11, CD247. CD7, CD81, CLSTN1, E2F6, MCM5, PARP1, PNPO, RASGRP1, RCC2, RPTOR, RUNX3 and ZAP70) had an AUROC to identify CTP2 higher than 0.95. Expression of these genes was synthesized into a transcriptomic score. As expected, the score was higher in CTP2 (Supplementary figure 7A), with an AUROC of 0.99 (95% CI 0.97 – 1) (Supplementary figure 7B). In a Cox-regression analysis including this transcriptomic score, age, sex and need for mechanical ventilation, the score was correlated to ICU discharge (HR 1.002 [1.000 – 1.003], p=0.012). Based on these results, a cut-off point of 250 in this score, aimed to include all CTP2 cases, was chosen.

Then, this transcriptomic score was calculated in an external cohort of 50 severe COVID-19 patients with publicly available blood gene expression in samples obtained at enrolment. After computing transcriptomic scores, 13 patients were classified as CTP1 and 37 as CTP2. Comparisons between these clusters are shown in Table 2. In spite of no significant differences in age, sex, APACHE-II or SOFA scores, patients assigned to CTP2 showed more ventilator-free days at day 28 of ICU stay, and the percentage of patients with zero ventilator-free days at day 28 was lower in CTP2. Deconvolution of peripheral blood transcriptomes in this validation cohort recapitulated some of the differences observed in the discovery cohort, including higher neutrophil counts and lower proportions of CD8+ T-cells in CTP1 (Supplementary Figure 8).

**Table 2.** Clinical data and outcomes in the validation cohort. APACHE-II: Acute Physiology and Chronic Health disease Classification System II. SOFA: Sequential Organ Failure Assessment. VFD: Ventilator-free days. P-values were calculated using a Wilcoxon test (quantitative data) or Chi-square test (proportions).

|  | CTP1 | CTP2 | p_value |
| --- | --- | --- | --- |
| Age (years) | 63 (55 - 73) | 64 (55 - 72) | 0.842 |
| Transcriptomic score | 216 (197 - 228) | 365 (311 - 470) | <0.001 |
| APACHE-II score | 23 (20 - 34) | 21 (14 - 25) | 0.097 |
| SOFA score | 7 (6 - 13) | 8 (6 - 10) | 0.35 |
| VFDs at day 28 | 0 (0 - 20) | 18 (2 - 28) | 0.016 |
| Zero VFDs at day 28 | 8 (62%) | 8 (22%) | 0.014 |

## Discussion

Our results show that unsupervised clustering of critically ill COVID-19 patients, using transcriptomic profiles from peripheral blood obtained at ICU admission, results in two groups with a differential immune response and outcome. In spite of no clinical differences at admission other than the absolute leukocyte count, the cluster of patients with an enriched interferon response shows lower ICU survival rates. Application of a cluster-specific score to an independent cohort confirmed this result. These findings suggest that there are specific COVID endotypes with different underlying immunopathogenesis and outcomes.

Clustering strategies have been proposed to identify different subgroups of critically ill patients with respiratory failure, that may help to personalize treatments. Two different phenotypes have been identified in several cohorts using clinical and laboratory data ^9,31^. A hyperinflammatory/reactive phenotype, characterized by higher concentrations of markers of acute inflammation, such as IL-6, IL-8, C-reactive protein, and tissue hypoxia has been linked to higher mortality rates and could specifically benefit from fluid restriction, higher PEEP levels or protective ventilation ^32^, in contraposition to the uninflamed phenotype. Of note, causes of ARDS were different between the two clusters, with a higher incidence of sepsis in the inflammatory/reactive group. Opposed to a syndromic approach, clustering within a specific disease such as COVID-19 using routine clinical data has yielded conflicting results. Whereas direct translation of the inflammatory/reactive framework to a single-center cohort has identified equivalent groups ^11^, other multicenter studies failed to identify COVID-19 subgroups at ICU admission ^12^.

In this setting, transcriptomic clustering may offer several advantages by including a large number of features for classification, reduced intervention times and absence of imputed or not available data. Bulk peripheral blood RNAseq has been used to identify relevant pathogenetic mechanisms in COVID-19, by comparing cases with different severity or against healthy controls^33–35^. Our approach selectively includes only severe cases and differs from these studies, revealing that critically-ill COVID-19 patients show two different transcriptomic profiles that include quantitative and qualitative differences in the regulation of the immune response to SARS-CoV-2 infection. Of note, these biological disparities occur despite no differences in clinical variables, suggesting that the identified clusters reflect endotypes with specific pathogenetic mechanisms and may outperform clinical diagnostic instruments. Survival in severe COVID-19 has been closely related to the host inflammatory response to viral infection. Genetic variants may result in different immune profiles with prognostic value. Polymorphisms in several chemokine receptors such as CCR1, CCR3, CCR9 or XCR1, or the interferon receptor (IFNAR2) have been linked to COVID-19 severity ^36,37^. Similarly, IFIH1 variants may result in an attenuated inflammation and a differential response to steroids ^8^. However, no genomic variants have been clearly linked to development of severe COVID-19 or death, and multiple variants may modify the immune response with diverging effects. This net result may be characterized using peripheral blood transcriptomes. Our unsupervised analysis of transcriptomic data (GSEA and cell deconvolution) revealed that patients with an interferon-driven response and a CD4+ T-lymphocyte profile show a worse ICU outcome compared to a cluster of patients with predominant B-cell and Treg activation. Previous studies focused in CD4+ T-cells and interferon signatures have yielded similar results in severe COVID-19 patients with marked proinflammatory responses ^38,39^. Although an enhanced interferon-mediated response may be detrimental, it must be noted that loss-of-function variants of genes from the interferon pathway (such as IRF7 or IFNAR1) or autoantibodies against interferons have been related to defective responses against SARS-CoV-2 and increased risk of severe COVID-19 ^40,41^ but not survival after ICU admission ^42^. Whereas T-cell activation is linked to interferon-dependent pathways, B-cell and regulatory T-cell activation seem to depend on upregulation of immune checkpoints, including BCL2 and members of the immunoglobulin and TNF superfamilies (IGHA1, IGHG1, CD27, LAG3 and TNFRSF18). Dysregulation of other immune checkpoints has been linked to mortality in COVID-19 patients ^43^. Collectively, these results highlight the need for a precise regulation of the inflammatory response after infection, avoiding not only hyper- or hypoinflammatory states, but also dysfunctional responses. Our results have several limitations. First, the sample size is reduced, so we cannot discard the existence of additional clusters with other underlying pathogenetic mechanisms, or that different clustering parameters or strategies may yield different results. However, unbiased p-values associated to the identified clusters were high, and the results confirmed in a validation cohort, thus suggesting a robust classification. Second, cell populations were estimated by deconvolution of the bulk transcriptome and should be confirmed using single cell RNA seq or flow cytometry. Finally, it is unclear if applied treatments can modify the expression of the genes used for clustering, although we did not observe differences in the prescribed treatments between groups. Moreover, it is unclear if therapeutic immunomodulation may impact this transcriptomic profile or, ultimately, outcomes.

In summary, our results show that transcriptomic clustering using peripheral blood RNA at ICU admission allows the identification of two groups of critically-ill COVID-19 with different immune profile and outcome. These findings could be useful for risk stratification of these patients and help to identify specific profiles that could benefit from personalized treatments aimed to modulate the inflammatory response or its consequences.

## Data Availability

All data and code used in the manuscript have been deposited in Git-Hub (https://github.com/Crit-Lab/COVID_clustering). FASTQ files with RNA and miRNA reads have been deposited in Gene Expression Omnibus (accession number GSE197259, available at https://www.ncbi.nlm.nih.gov/geo/query/acc.cgi?acc=GSE197259).

https://www.ncbi.nlm.nih.gov/geo/query/acc.cgi?acc=GSE197259

## Acknowledgements

The authors want to thank all the personnel at the participating ICUs and laboratories for their support during the development of the study. This work is supported by Centro de Investigación Biomédica en Red (CIBER)-Enfermedades Respiratorias (CB17/06/00021), Instituto de Salud Carlos III (grants PI20/01360 and PI21/01592, FEDER funds) and Fundació La Marató de TV3 (413/C/2021). CLM is supported by Ministerio de Universidades, Spain (FPU18/02965). RRG is supported by a grant from Instituto de Salud Carlos III (CM20/0083). PMV is supported by a grant from Instituto de Salud Carlos III (FI21/00168). Instituto Universitario de Oncología del Principado de Asturias is supported by Fundación Liberbank. AD is supported by the Comunidad de Madrid and European Regional Development Fund (REACT EU Program “FACINGLCOVID-CM”).

## Author contributions

Study design and supervision: GMA, LAR. Sample acquisition and processing: MFR. RNA Sequencing: CLM, PMV, ILA, JGdO, HGP, ECL, IC, EC. MiRNA sequencing: CLM, AD, LACh. Patient inclusion, follow-up, and data collection: ESdR, RRG, DP, GMA, LAR. Data analysis: CLM, PMV, LACh, GMA, LAR. Results discussion: CLM, PMV, ILA, IC, JRC, AD, EC, GMA, LAR. Manuscript writing: CLM, GMA, LAR. Manuscript review and editing: All authors.

## Competing interests

The authors declare no competing interests.

## Corresponding authors

Correspondence and requests to Laura Amado-Rodríguez or Guillermo M Albaiceta.

## Notes

### Competing Interest Statement

The authors have declared no competing interest.

### Funding Statement

This work is supported by Centro de Investigacion Biomedica en Red (CIBER)-Enfermedades Respiratorias (CB17/06/00021), Instituto de Salud Carlos III (grants PI20/01360 and PI21/01592, FEDER funds) and Fundacion La Marato de TV3 (413/C/2021). CLM is supported by Ministerio de Universidades, Spain (FPU18/02965). RRG is supported by a grant from Instituto de Salud Carlos III (CM20/0083). PMV is supported by a grant from Instituto de Salud Carlos III (FI21/00168). Instituto Universitario de Oncologia del Principado de Asturias is supported by Fundacion Liberbank. AD is supported by the Comunidad de Madrid and European Regional Development Fund (REACT EU Program FACINGLCOVID-CM).

### Author Declarations

This prospective observational study was reviewed and approved by the regional ethics committee (Comite de Etica de la Investigacion Clinica del Principado de Asturias, ref 2020.188). Informed consent was obtained from each patient next of kin.

