## Supplementary material for "Transcriptomic clustering of critically ill COVID-19 patients": Online supplement

### Online data supplement

Supplementary figure 1. Hierarchical clustering tree showing the p values for each cluster, calculated using the pvclust package for R. AU (red): Approximately unbiased p-values. BP (blue): Bootstrap probability.

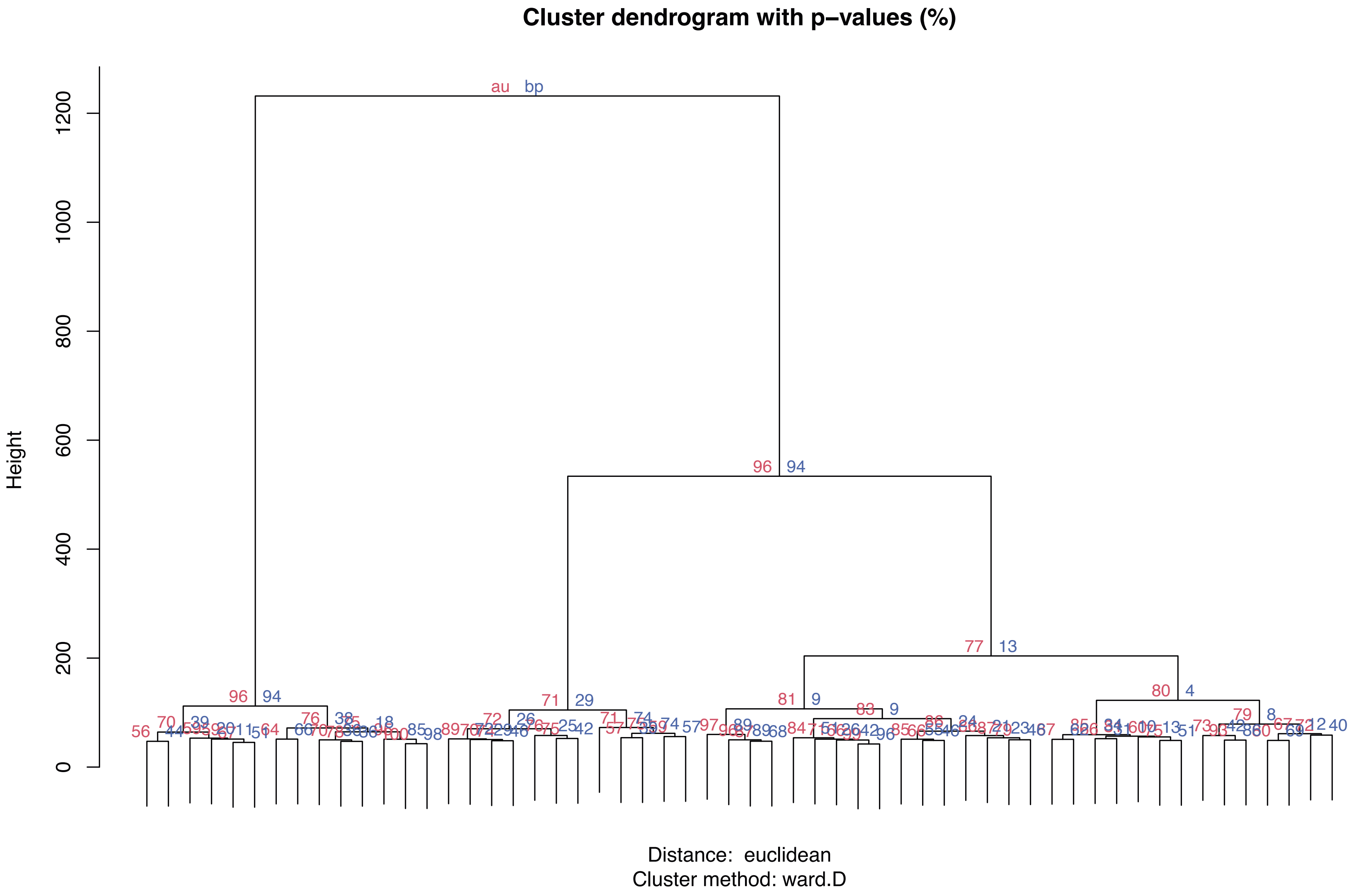

**Supplementary figure 2.** Heatmap showing differences in expression between COVID transcriptomic profiles (CTP) in the 1727 genes used for clustering.

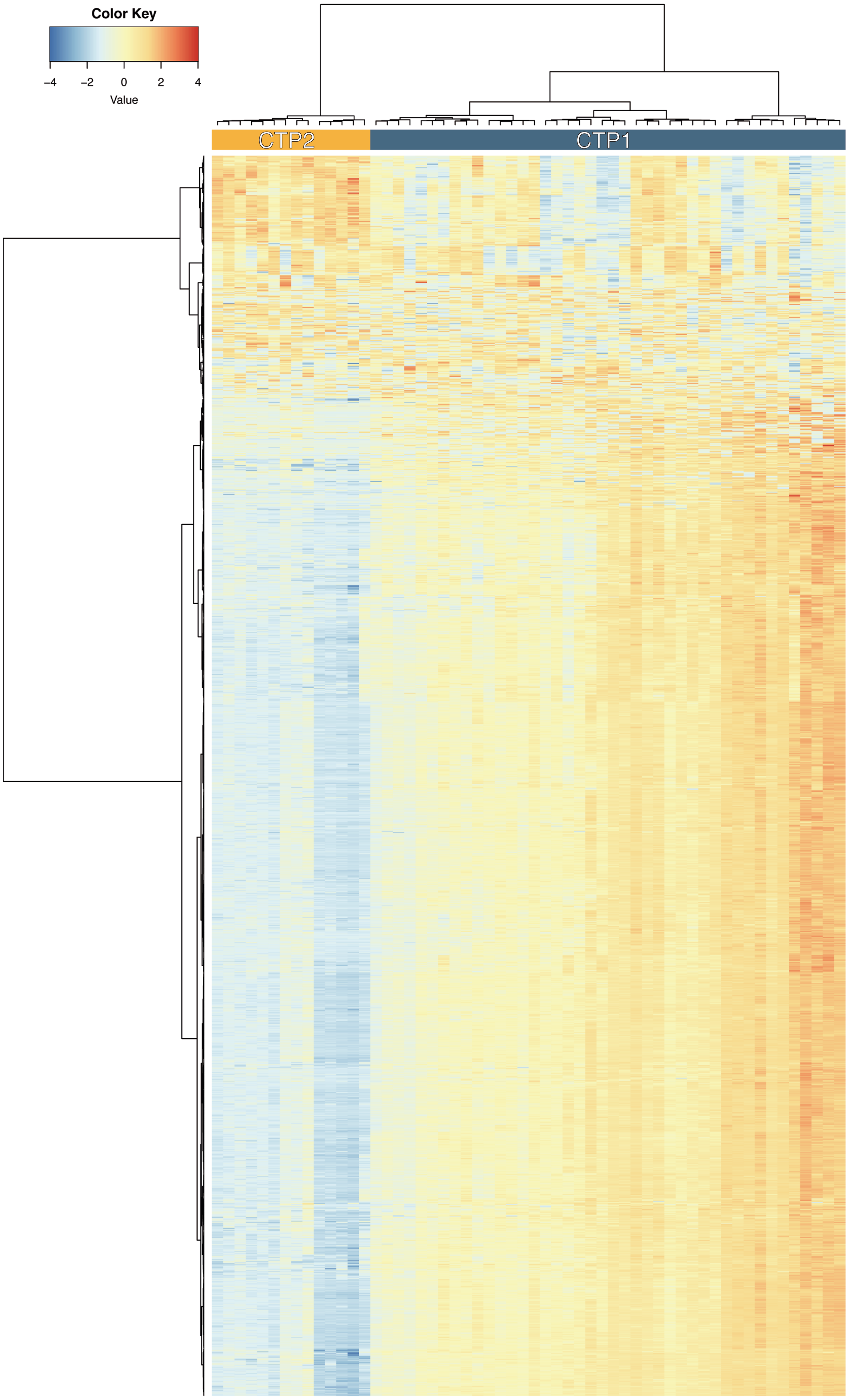

**Supplementary figure 3.** Biological processes with significant differences in gene enrichment between COVID transcriptomic profiles (CTP).

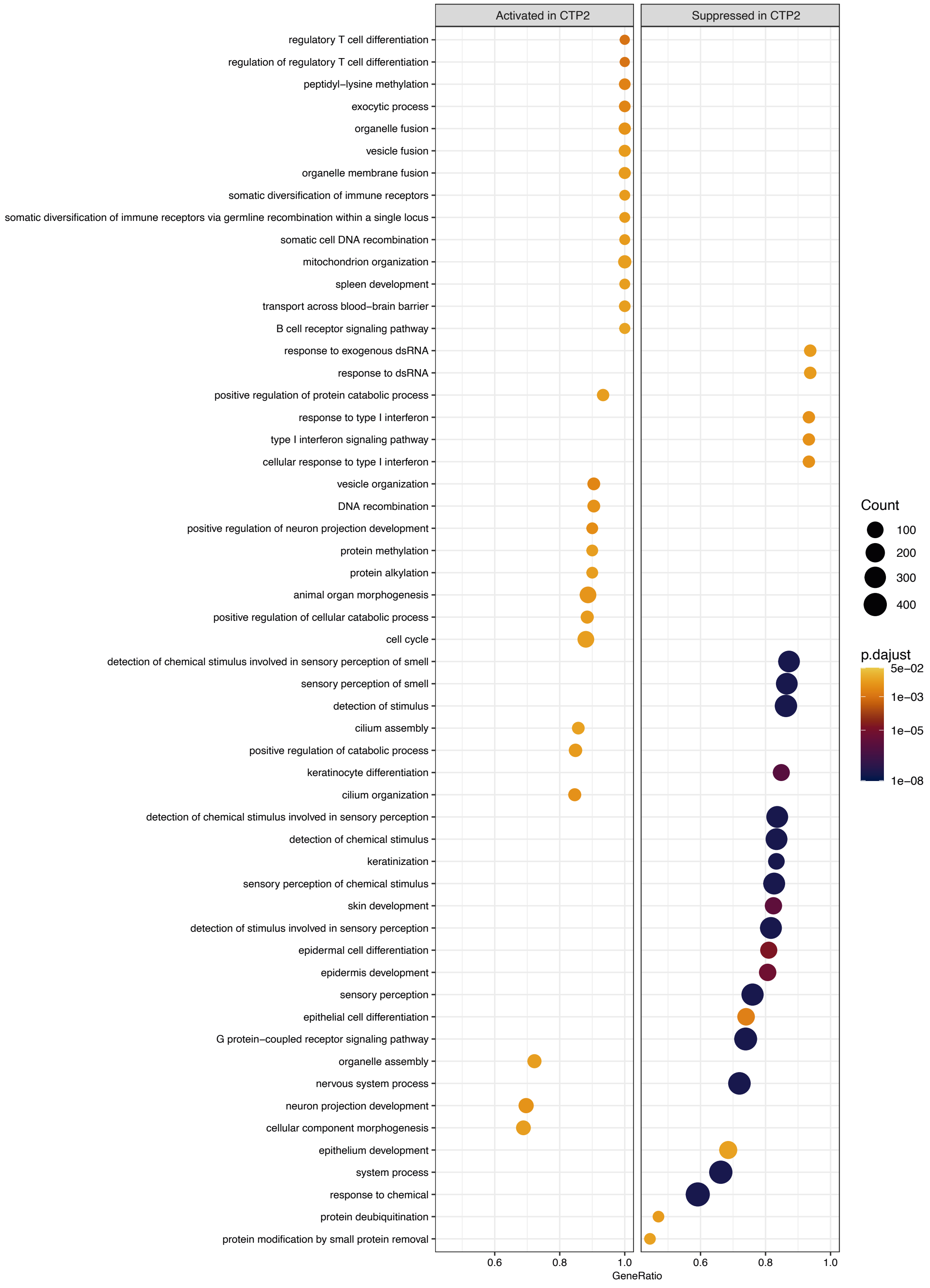

**Supplementary figure 3 (cont).** Biological processes with significant differences in gene enrichment between COVID transcriptomic profiles (CTP).

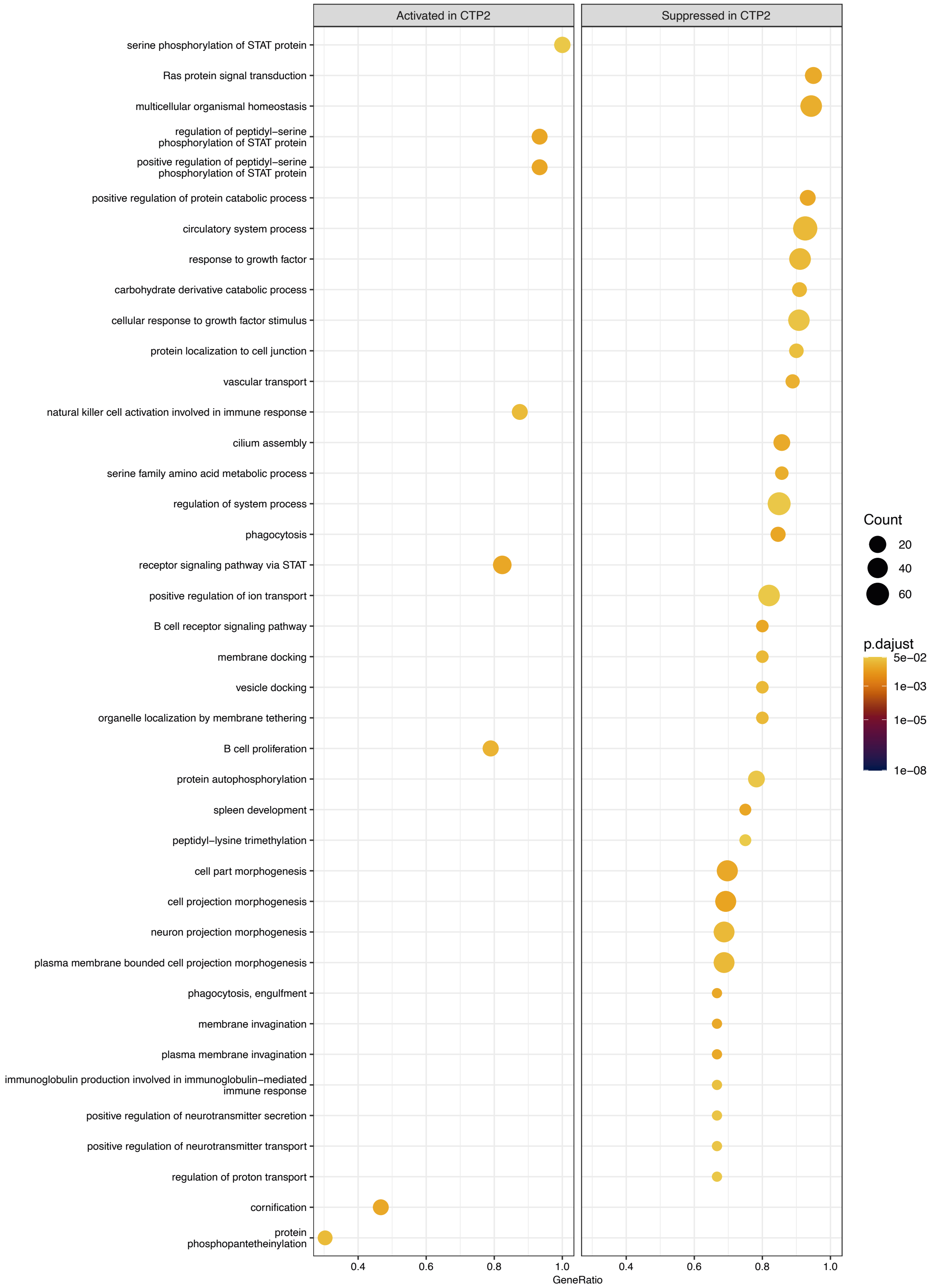

**Supplementary figure 4.** Correlations between genes in each COVID transcriptomic profile (CTP).

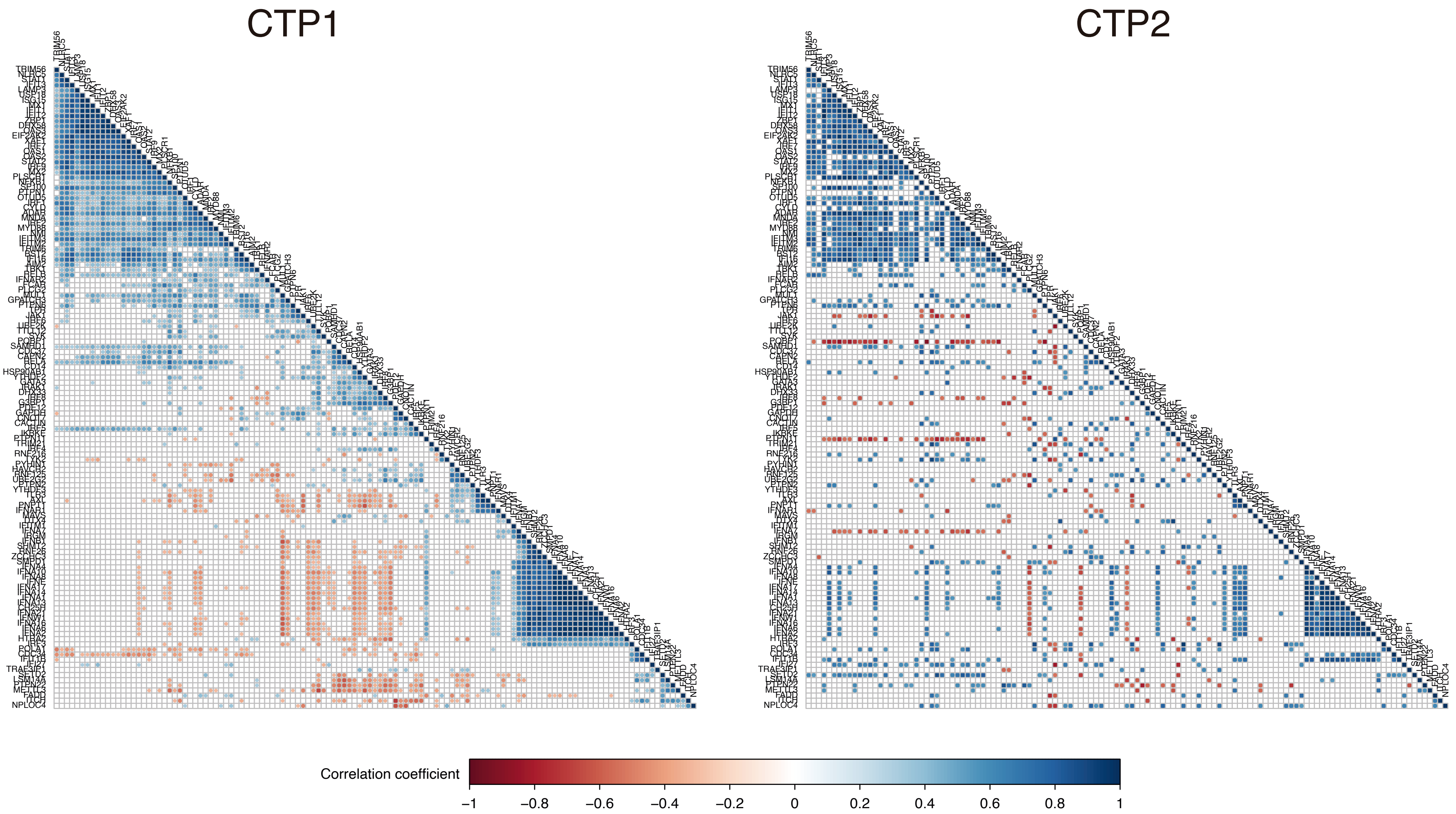

**Supplementary figure 5.** Differences in estimated cell populations between clusters. CTP: COVID transcriptomic profile. Points represent individual patient data. In boxplots, bold line represents the median, lower and upper hinges correspond to the first and third quartiles (the 25th and 75th percentiles) and upper and lower whiskers extend from the hinge to the largest or smallest value no further than 1.5 times the interquartile range. P-values were calculated using a two-tailed Wilcoxon test.

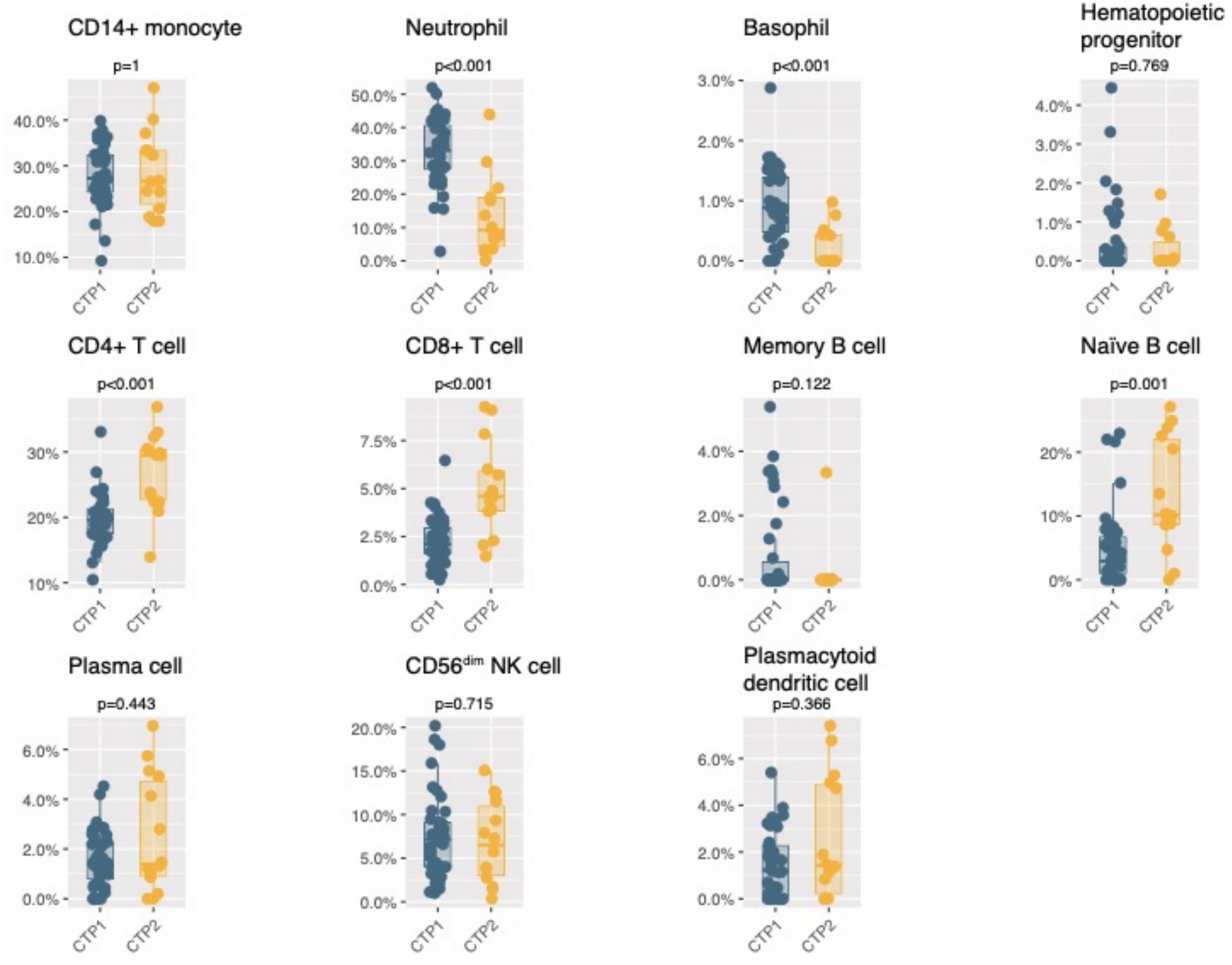

**Supplementary figure 6.** A: MicroRNA/gene network showing predicted upregulated miRNAs related to downregulated genes in patients with COVID transcriptomic profile (CTP) 2. B-M: Normalized counts of each miRNA. Points represent individual patient data. In boxplots, bold line represents the median, lower and upper hinges correspond to the first and third quartiles (the 25th and 75th percentiles) and upper and lower whiskers extend from the hinge to the largest or smallest value no further than 1.5 times the interquartile range. All p values (Wilcoxon test) were higher than 0.05.

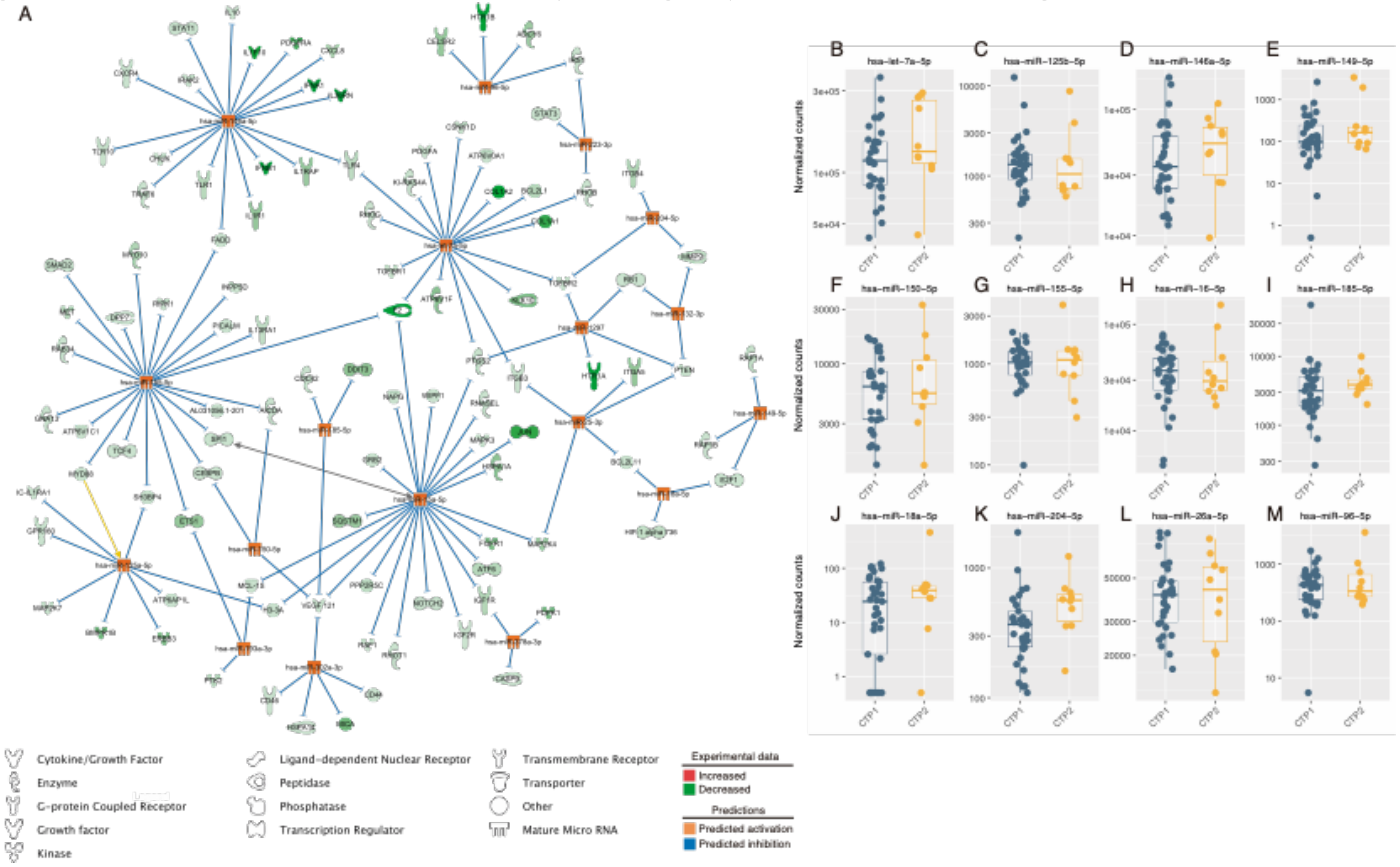

**Supplementary figure 7**. A: Transcriptomic scores calculated in the training sets. B: ROC curve corresponding to the diagnostic accuracy of the transcriptomic score to identify COVID transcriptomic profiles.

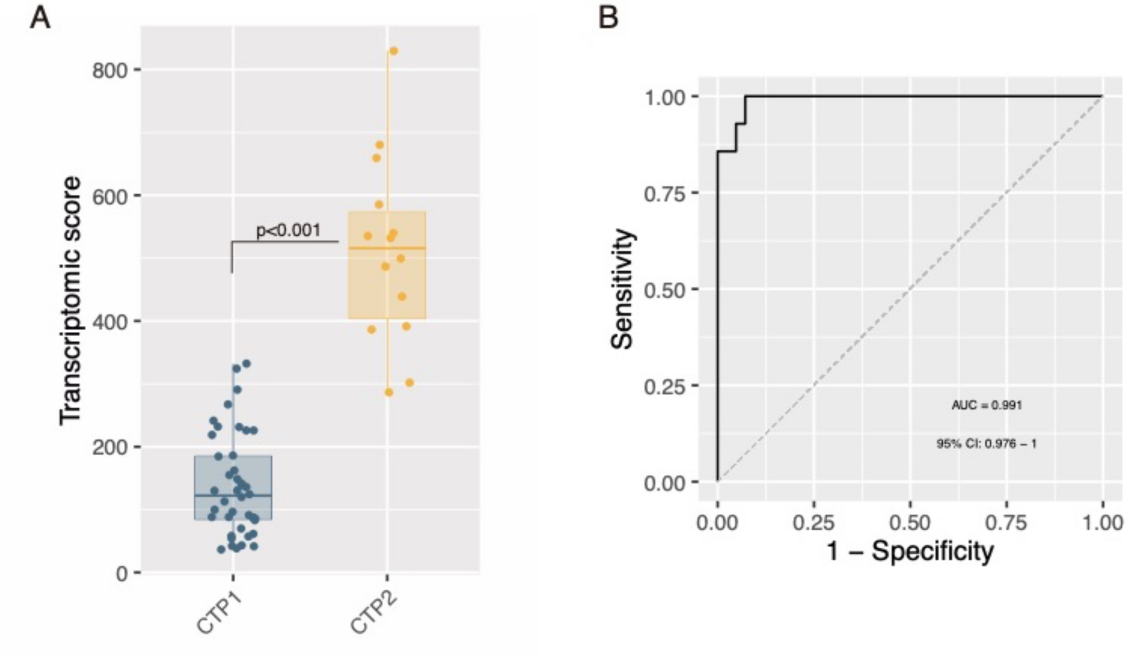

**Supplementary figure 8.** Estimated cell populations according to COVID transcriptomic profiles (CTP) in the validation cohort. Points represent individual patient data. In boxplots, bold line represents the median, lower and upper hinges correspond to the first and third quartiles (the 25th and 75th percentiles) and upper and lower whiskers extend from the hinge to the largest or smallest value no further than 1.5 times the interquartile range. P-values were calculated using a two-tailed Wilcoxon test.

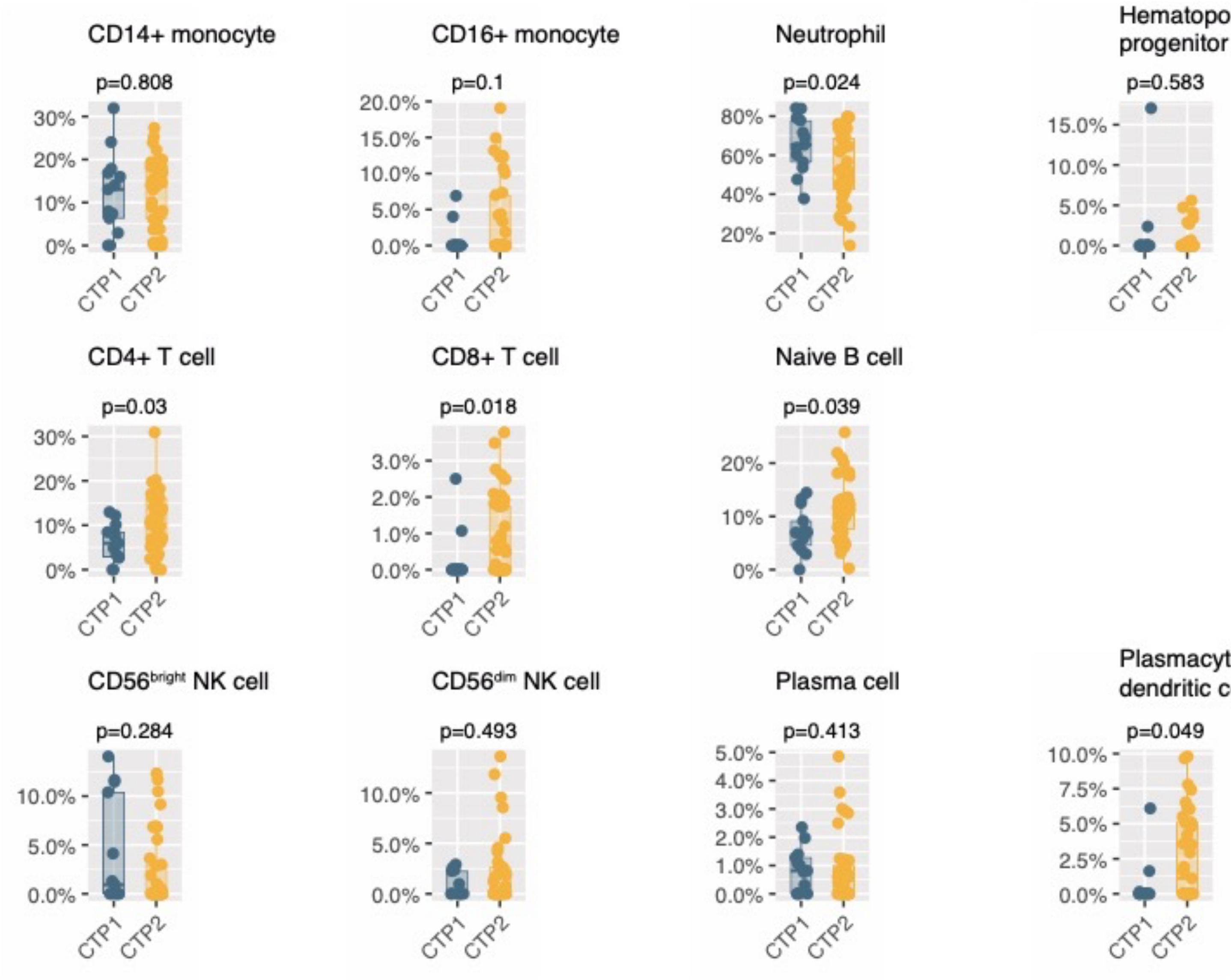
